# An inflatable ‘finger-lock’ for stabilizing nailfold capillary videos and modulating blood flow velocities

**DOI:** 10.1101/2025.02.24.25322798

**Authors:** Marcus L. Forst, Gabriela Rincon, Juliette H. Levy, David N Cornfield, Stephen R Quake

## Abstract

High-resolution video capillaroscopy shows promise as a non-invasive method to directly observe blood cells as they flow through and interact with the microvasculature. When imaging microvasculature in the nailfold, the inherent shaking of the fingers requires physical stabilization for image noise reduction. However, any external force applied to the fingers will affect the rate of blood flow. We designed an inflatable ‘finger-lock’ for nailfold capillaroscopy, which stabilizes the finger against a coverslip with constant pressure. Testing 72 participants, we demonstrated that increasing ‘finger-lock’ pressure improves video stability and decreases the velocity of capillary blood cells. This shows that capillary blood cell velocity measurements should monitor and control external pressure. Our work introduces a method to perturb local capillary blood flow and measure the microvascular response, enabling further studies investigating how person-specific factors (e.g. age, disease) impact vascular health.

## 1 Introduction

Capillaries connect the cardiovascular system to our tissues, transferring essential oxygen and nutrients across their permeable membranes. Although they are the smallest vessels in the body, photographing superficial capillaries in the nailfold is straightforward–moisturizing the skin allows visible light to penetrate deep enough (500-2500 *µ*m[1, 2]) into the skin for adequate contrast. Images of superficial capillaries in the nailfold (depth of 150-400 *µ*m)[3] have long been used to diagnose and grade vascular diseases like Raynaud’s Syndrome[4] and Systemic Sclerosis [5–8] by analyzing their tortuousity[4, 6, 9, 10], uniformity[11], density[12, 13], and size[5, 7, 8, 11, 14, 15]. Other methods such as skin temperature testing[16], capillary refill[17, 18], venous occlusion plethysmography (VOP)[19, 20], and laser-Doppler flowmetry (LDF)[12, 18, 21–27] have been used to measure bulk capillary flow dynamics but traditional capillaroscopes have lacked the resolution and framerate needed to distinguish individual cells and analyze their movement through capillaries. Recent advances in cameras and computer vision have enabled high spatiotemporal-resolution videos of capillary blood flow, facilitating in-vivo single-cell tracking[28–31], white blood cell (WBC) counting and classification[28–30, 32–36], rolling WBC visualization[31, 37, 38], and pulse-wave velocity measurement[39]. However, the high magnification required to image blood cells significantly amplifies any small movement, adding spatial noise and hindering effective data analysis. Every capillaroscope has a method for stabilization; retinal imaging requires pupil dilation and head vibration isolation[40] and oral mucosa imaging requires inserting a vacuum or clamp into the mouth[28, 31]. Nailfold imaging requires placing hands on a table[41], in a fingerholder[32, 35, 42], or pressing the capillaroscope directly on the finger[43]. But, pressing on capillaries for stability—whether by pressing, clamping, or holding—inevitably modifies blood flow. To ensure meaningful results, we must either quantify the pressure applied and extrapolate back to determine the unperturbed velocity or maintain a consistent pressure across measurements. Without this, the stabilization method itself becomes a confounding variable. In this work, we address finger shaking by developing an inflatable ‘finger-lock’ to stabilize the finger with constant pressure against a coverslip. This improves video quality and enables automatic video processing to measure blood cell velocities. We demonstrate that increasing external pressure slows capillary blood flow, confirming that we must measure blood cell velocities at constant pressure and extrapolate to estimate blood cell velocities at zero pressure. Further, by modulating external pressure, our finger-lock shows promise for assessing capillary reactivity, revealing how the microvasculature responds to mechanical perturbation.

## 2 Design Overview

### 2.1 Optical Setup

Nailfold capillaries lie roughly 150 *µ*m underneath the surface, meaning that much of the light entering the finger absorbs or scatters away[44–46]. To preserve as much light as possible, we designed our optical setup with minimal components, aligning an objective, a tube lens, and a camera in a bright field configuration (Fig. 1). We chose a 0.25 NA 10x achromatic objective to obtain sufficient resolution for individual red blood cells while maintaining a large enough field of view to locate easily visible capillaries (Fig. 2 a-b). Our objective had a large enough working distance to illuminate the sample from the side, rather than through the objective, enabling us to limit light absorption and avoid back reflections to the camera. Moreover, the 0.25 numerical aperture reduces the depth of field, reducing interference from melanin in our images, thus enabling our microscope to image capillaries effectively on participants of all skin tones (Fig. 3). To minimize aberrations and maximize photon capture, we focused light onto our camera using a 2” diameter, 150 mm focal length achromatic tube lens. We used a monochromatic Basler Ace acA1440-220um USB camera with Basler Pylon software for ease of use and high frame rate. With this setup, we achieved a resolution of at least 645.1 lpm or 1.55 *µ*m (Fig. 4 d-f), a magnification of 8.418x, and a maximum frame rate of 227.8 fps.

**Fig. 1:**
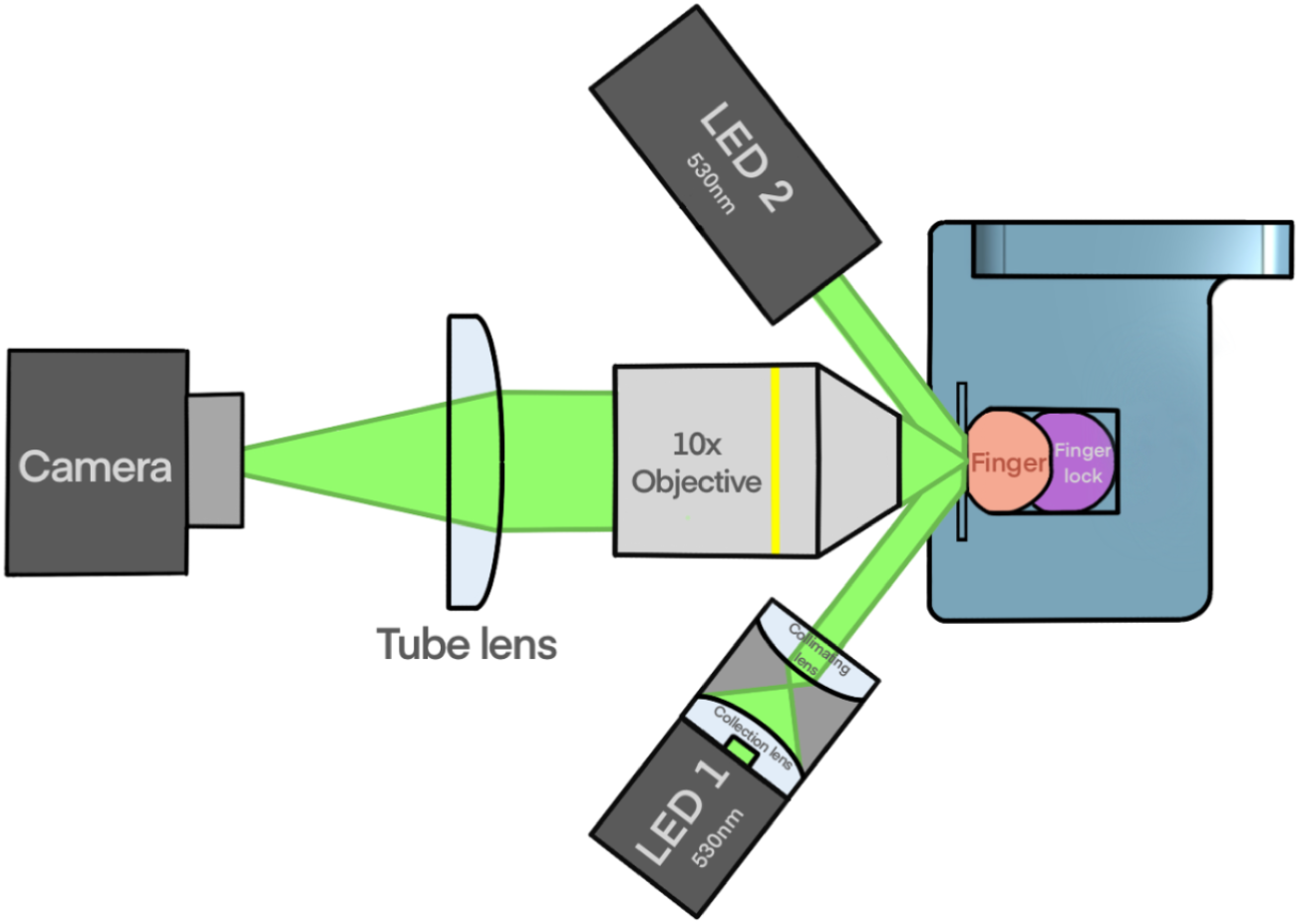
Top-down schematic of the capillaroscope. We built a bright-field capillaroscope with a 10x objective, 150nm tube lens, and 227.8 fps Basler Ace camera. Two 530 nm Thorlabs M530L4 LEDs, maximally defocused in the Kohler configuration, obliquely illuminate the finger. A 3D-printed fingerholder holds the finger against a glass coverslip with constant pressure using an inflated nitrile glove, or ‘finger-lock’. LED light is collected by an aspheric lens and focused directly before a second, defocusing lens. The resulting beam is directed to the finger at an angle of 30 degrees.

**Fig. 2:**
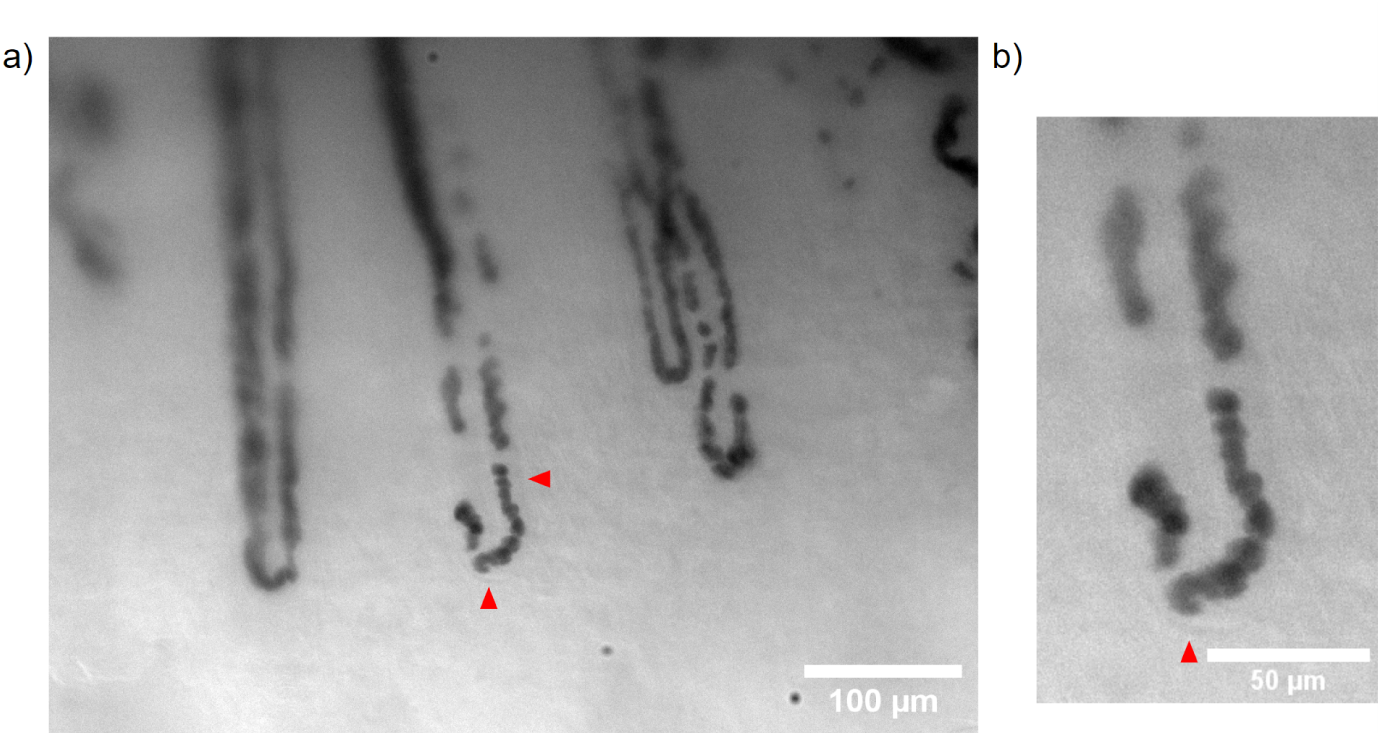
Capillary image shows individual red blood cells. a) Capillary image showing hairpin-shaped nailfold capillaries filled with individually discernable red blood cells. Some of the capillary wall is visible, shown by red triangles. b) Zoomed-in capillary showing red blood cells flowing around the capillary loop, squishing together as they squeeze past the hairpin.

**Fig. 3:**
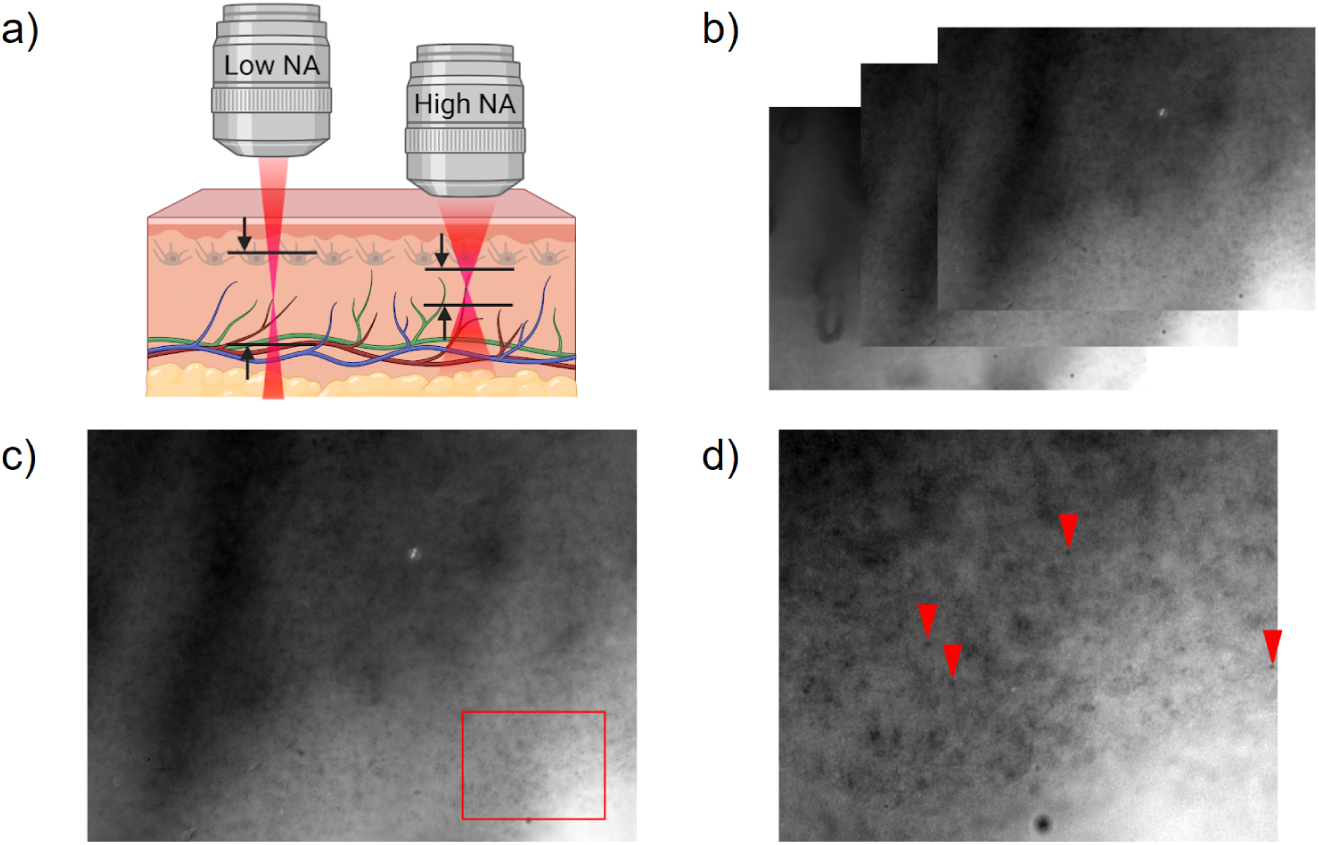
High numerical aperture (NA) objective reduces depth of field and thus removes interference from melanin and melanocytes. a) Schematic showing how reduced depth of field limits the interference of melanin and melanocytes. The black lines and arrows illustrate the depth of field for each respective objective. b) A stack of images at different image planes shows melanin closer to the surface and capillaries further down. c) An image of a melanin-filled layer. We can see the shadows of the capillaries underneath. d) The zoomed-in image inset shows melanin (red arrows show examples).

**Fig. 4:**
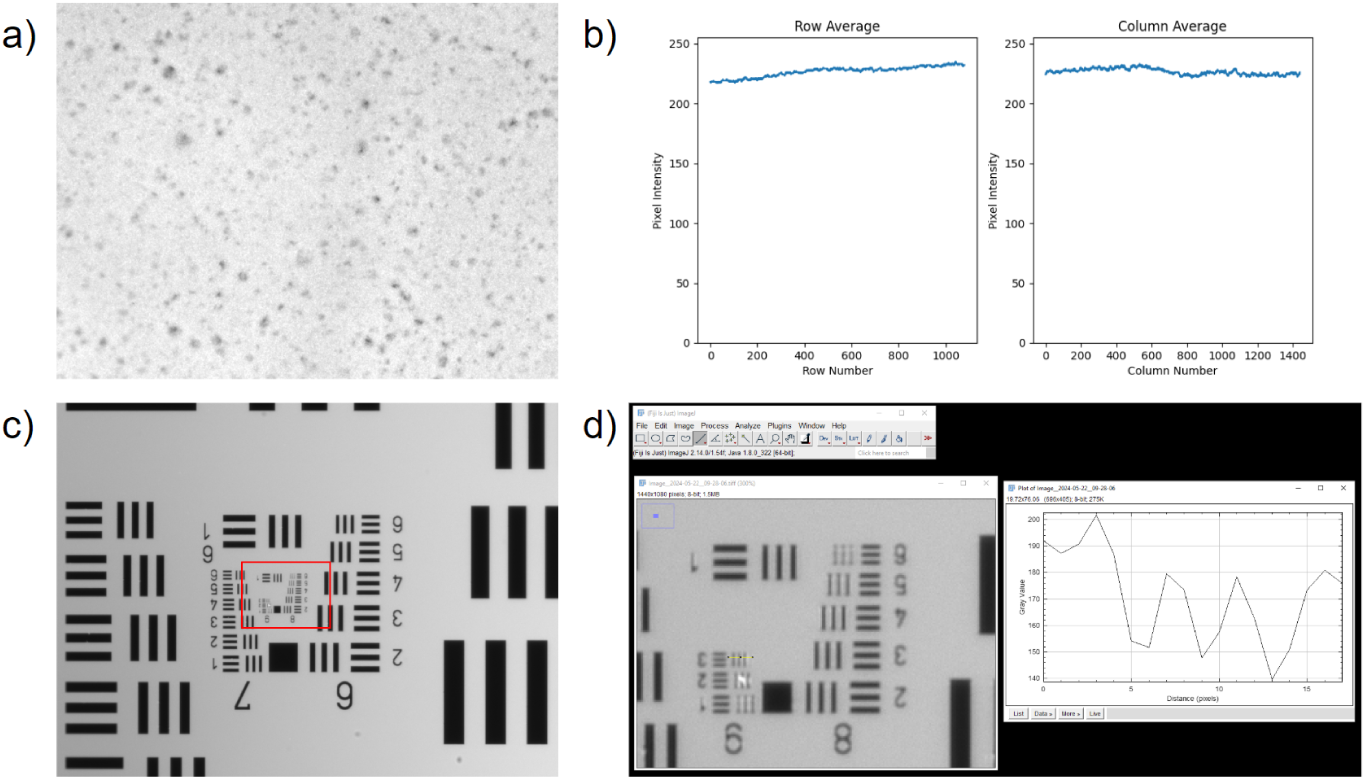
Illumination and resolution benchmarking show resolution of better than 645.1 lpm (1.55 *µ*m). a) The frosted end of a glass slide, used to evaluate illumination, as seen in the microscope. b) Average illumination of rows (vertical line illumination average). The x axis is the row index, the y axis is the average intensity. c) Average illumination of columns (horizontal line illumination average). The x axis is the column index, the y axis is the average intensity. d) 1951 USAF resolution target shows maximum resolution of better than 645.1 lpm (1.55 *µ*m).

### 2.2 Oblique Kohler Illumination

We illuminated the finger with two beams of green 530 nm light from a pair of Thorlabs M530L4 LEDs oriented at an angle of 30 degrees with the nailfold to direct surface reflections away from the camera. We used the Kohler configuration to evenly illuminate the nailfold without LED artifacts (Fig. 4 a-c); An aspheric lens collected light from the diode before another lens maximally defocused it in the image plane (Fig. 1). We chose green 530 nm light because of its strong absorption by hemoglobin[47], which gave significantly higher RMS contrast and Weber contrast compared to a Thorlabs MCWHL6 white broadband LED (Fig 5 a-b). Using 530 nm light also improves robustness with different skin tones because the ratio of hemoglobin absorption to melanin absorption is higher than at other wavelengths [47, 48]. Like Bourquard et al, we use two obliquely-angled LEDs for illumination[32, 49]. The second LED increased the signal-to-noise ratio (SNR), minimized shadows and phase contrast, and enabled higher-frame rate, full-contrast videos (Fig. 5 b-d). Though phase contrast has been effectively used for WBC classification and counting in the tongue and lip[28], we prioritized light and framerate to enable our microscope to accurately measure blood cell velocities.

**Fig. 5:**
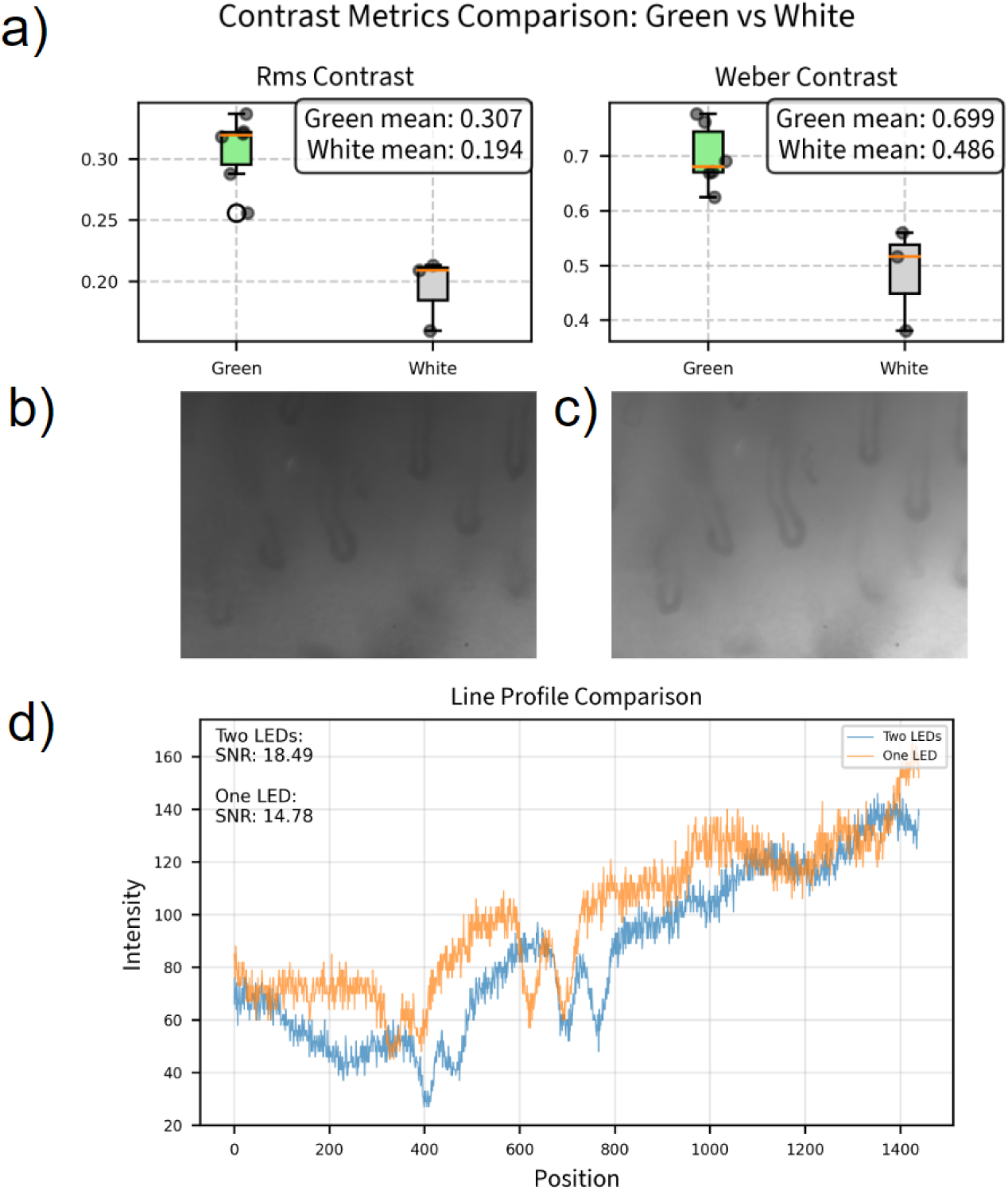
530nm LED and symmetric illumination improve RBC contrast. a) RMS and Weber contrast comparison between images taken with 530 nm Thorlabs M530L4 LED and Thorlabs MCWHL6 white broadband LED. Using the green LED improves contrast. b) Image of capillaries using single LED with 5074 *µ*s exposure time. c) Image of capillaries using two LEDs with 5074 *µ*s exposure time. d) Line profile of the center row shows that adding a second LED improves signal-to-noise ratio (SNR) from 14.78 to 18.49 and contrast from 49.4 to 52.8.

### 2.3 3D-Printed Finger Holder

To stabilize the finger, we first reduced arm and wrist motion using an armrest and wristrest (Fig 6 a). We designed a 3D-printed fingerholder with a rectangular cutout to stop the finger from moving side-to-side. Figure 6 b shows a 17.3 mm (ring size 7) cutout because that is the size of an average finger, but we made versions with other ring sizes. We extruded a 300 degree cone for oblique illumination of the finger (Fig. 6 c-d), reinforced on the inside of the chamber by curving filets performing the double benefit of stabilizing the finger. The front of the fingerholder includes a slot for the coverslip (Fig. 6 b), which ends at the bottom of the illumination cutout, hanging slightly over the cuticle but stopping before the fingernail to create a seal with the nailfold (Fig. 6 d). This seal holds the mineral oil–which moisturizes the finger and enables imaging through skin–by spreading it evenly against the glass slide via capillary action, dramatically improving resolution without a mineral oil bath. Another cutout at the bottom of the channel accommodates fingernails extending beyond the finger when the fingertip is compressed (Fig. 6 d). Behind the finger, we cut a hole large enough to snugly insert a pneumatic air tube covered by a nitrile glove–our ‘finger-lock’. The hole was placed 1.5 cm behind that finger so the finger-lock touched the entire finger when inflated to 0.2 psi and above.

**Fig. 6:**
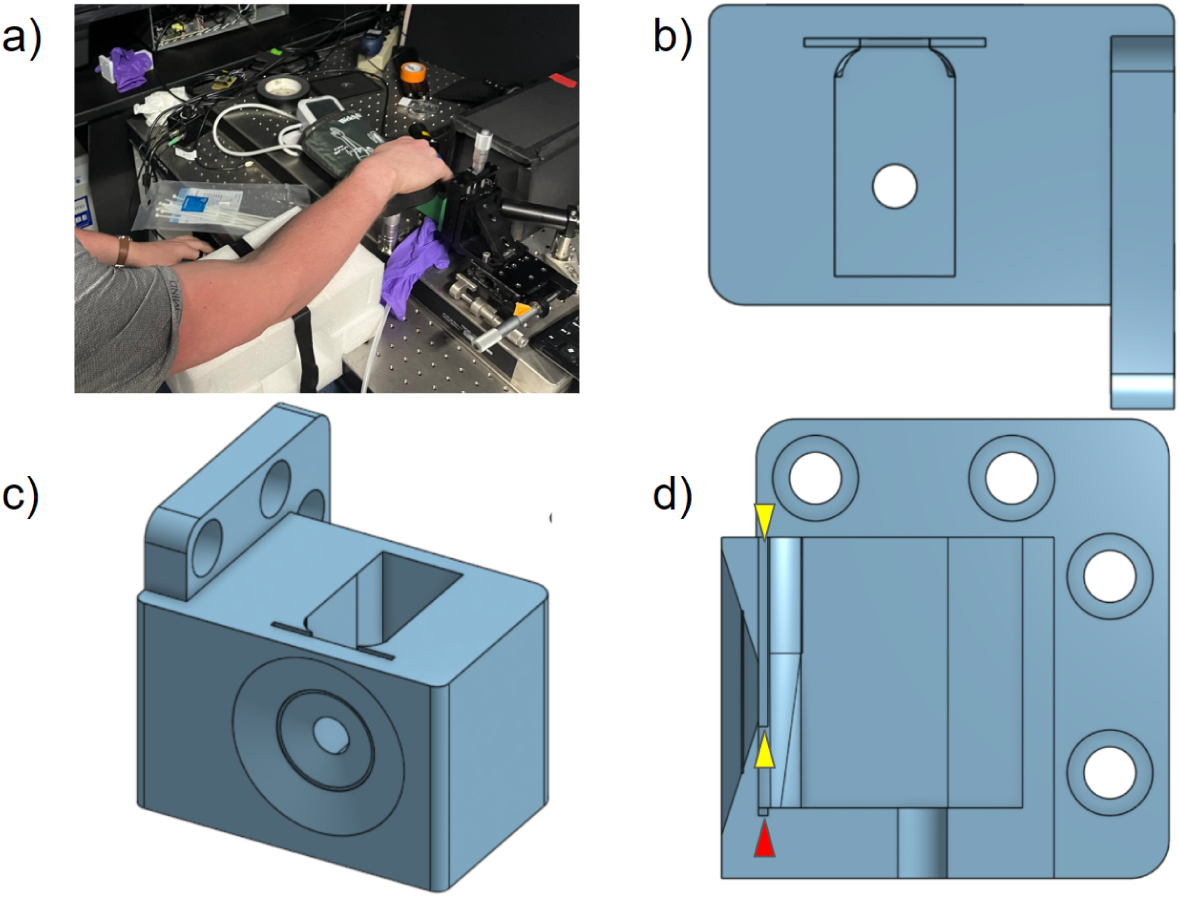
Armrest, wristrest, and fingerholder design. a) The armrest and wristrest stabilize the arm. b) A top-down view of fingerholder shows the channel for the finger, a coverslip slit, curving fillets for stabilization of the finger and coverslip, and a hole for the finger-lock. c) A front-angle view of fingerholder shows the 30-degree cutout for oblique illumination. d) Cutout side-view of fingerholder. The coverslip slit length is shown with two yellow arrows, note that it ends at the bottom of the illumination hole. The fingernail slit is shown with a red arrow and could be made larger for people with extended nails.

### 2.4 Inflatable “Finger-Lock”

The ‘finger-lock’ is created by inflating the middle finger of a medium-sized nitrile glove with compressed air. We controlled the pressure using an Enfield TR-010-V-EX pressure regulator and SP3-GB35-M12-U01 pressure sensor with 350 ml air reservoir. Pressure regulators have a robust analog feedback loop that automatically keeps the balloon’s pressure constant. To avoid exceeding the regulator lockup pressure, two step-down regulators brought the air pressure from the compressed air source (constant flow) to 100 psi and then down to 5 psi (Fig. 7 c). We chose 5 psi because 4 psi was the highest pressure needed to stop blood flow in the participants we measured. The Enfield regulator requires the inlet pressure to be 1 psi higher than the highest outlet pressure. The finger-lock fills the cavity and makes full contact with the finger, applying constant pressure to hold the finger steadily against a coverslip (Fig. 7 a). We used the Enfield microcontroller software to adjust the pressure up and down by 0.2 psi increments and image the results. To standardize our procedure, we took videos for every participant in increments of 0.2 psi, starting at 0.2 psi, increasing to 1.2 psi, and decreasing back down to 0.2 psi (Fig. 7 b). For participants requiring more than 1.2 psi to stop blood flow, we increased pressure until their flow stopped, and then took a video.

**Fig. 7:**
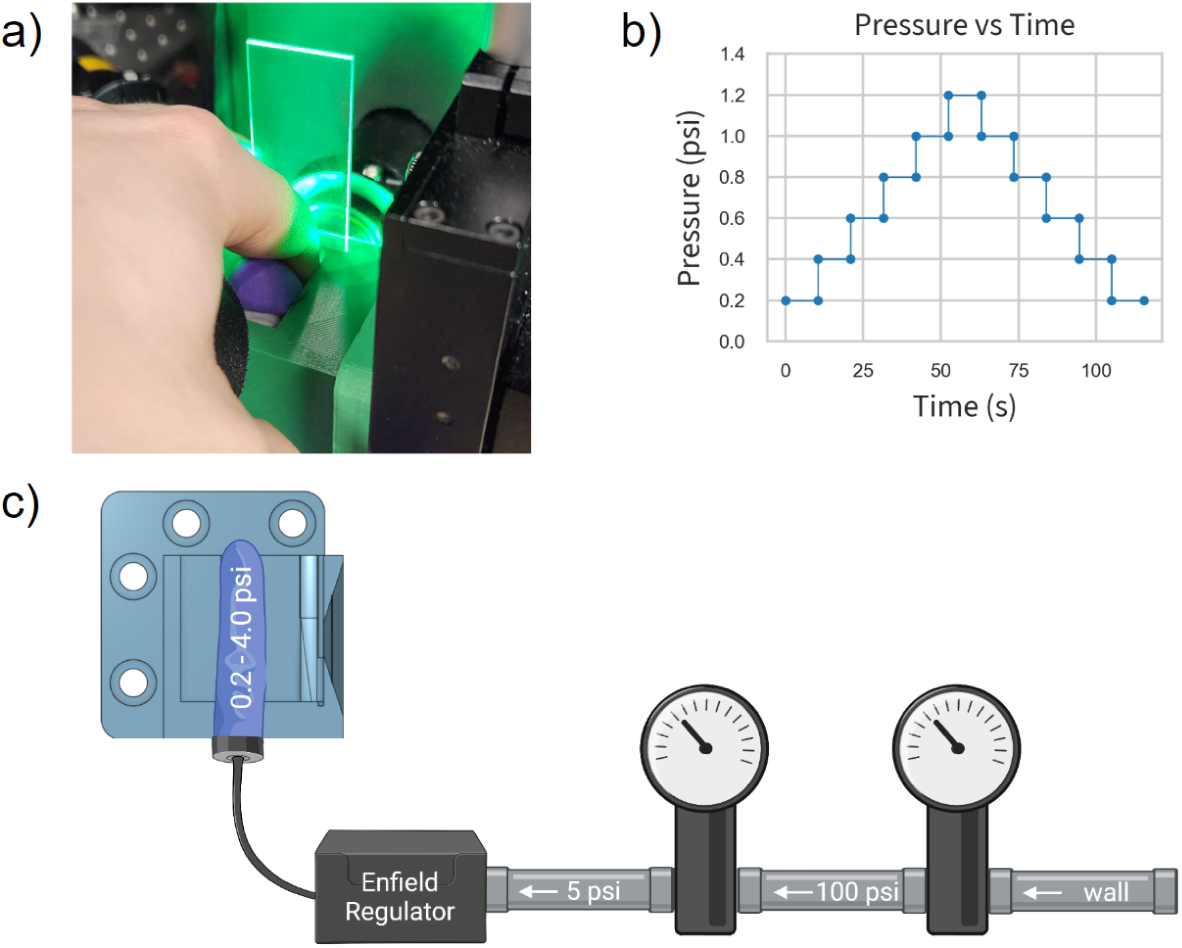
Finger-lock design and usage. a) Inflated “finger-lock” holds finger against coverslip. b) The pressure ramp and descent are controlled by the Enfield pressure regulator. Each video was 625 frames, taken at 113.9 fps. After the video was taken, the camera buffered for 5 seconds. During this time, we increased the pressure by 0.2 psi and refocused the camera. Hence, we took a 5.49-second video every 10.49 total seconds. c) Two step-down pressure regulators reduce the pressure to 100 psi and then to 5 psi at the Enfield regulator inlet. The Enfield regulator then controls the pressure inside a nitrile glove “finger-lock” to hold the finger against a coverslip.

### 2.5 Velocity Analysis Pipeline

To measure velocities, we created and analyzed spatiotemporal plots–called kymographs–and measured the slopes of blood cell trajectories to get an average blood cell velocity for each capillary in each video.

#### 2.5.1 Finger Location Selection

We primarily used the ring and pinky fingers for videos because their capillaries are closer to the surface, giving better images. We looked for locations near the cuticle where multiple capillaries were exposed and flowing. Specifically, we used capillaries at the top of the cuticle, where the finger presses on the glass slide. Once we found a flowing capillary, we took a series of videos, choosing one capillary to focus on as we increased the pressure. We adjusted the field of view to keep the chosen capillary in frame. We would select the second location based on the following trends: Ring fingers had larger but deeper capillaries than pinky fingers. If our first location had big capillaries that were too deep, we would check the pinky to see if we could trade size for clarity. If ring finger capillaries were too small, we would try the pointer and then middle fingers for bigger capillaries. Most often, the ring finger was the best choice.

#### 2.5.2 Data Pipeline

Participant videos were grouped by finger location and analyzed to extract velocity data of individual capillaries. To sharpen capillary edges for segmentation, we averaged frames by taking the median of video stacks stabilized using the “MOCO” ImageJ plugin[50] with a downsample value of 2 (Fig 8 a). We checked each video to ensure the videos were stable and sliced videos if a portion of the video was out of focus or incorrectly stabilized.

**Fig. 8:**
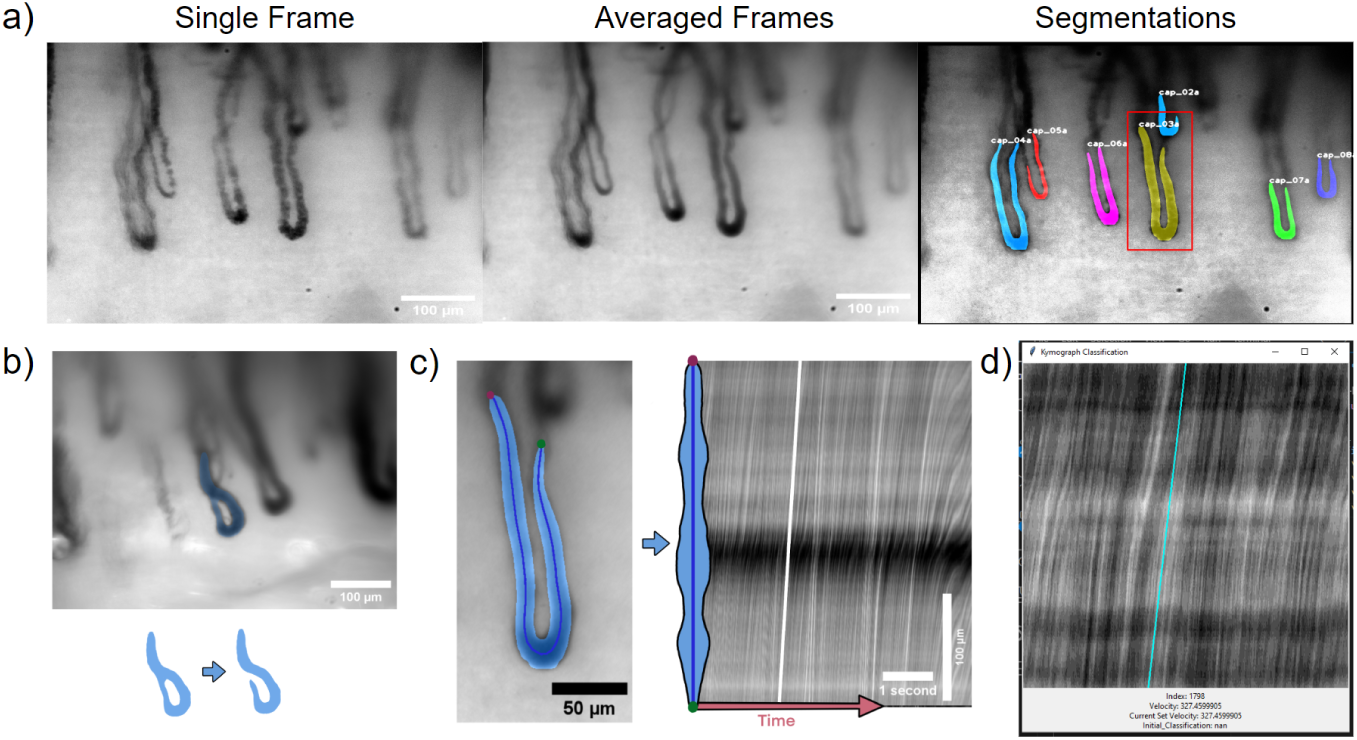
Image processing pipeline creates kymographs to calculate velocities. a) Averaging all frames in a stabilized video sharpens capillary boundaries for segmentation by hasty.ai algorithm. The third image shows multiple capillaries identified in the same video. b) Our centerline pruning algorithm cannot prune loops so we manually edit segmentations to remove loops. c) For each segmentation, we find the centerline and plot each centerline pixel with respect to time, creating a kymograph with time on the x-axis and centerline position on the y-axis. Diagonal lines show blood cell packet trajectories. We calculate the average trajectory slope, seen here in a white line, to measure the average blood cell velocity. d) We created a GUI to check velocity correctness and manually reclassify velocities into specific velocity ranges if incorrect.

#### 2.5.3 Segmentation

We segmented capillaries using a custom-trained “hasty.ai” [51] neural net, editing segmentations to remove loops and overlapping capillaries (Fig. 8 a-b).

#### 2.5.4 Centerlines

To calculate the centerline of our capillary segmentations, we used the FilFinder skeletonize algorithm [52] and pruned centerlines with a branch threshold of 40 pixels (16.4 *µ*m), a minimum capillary length of 50 pixels (20.5 *µ*m), and a prune criterion of ‘length’. For the pruning to work correctly, we required segmentations without loops.

#### 2.5.5 Kymographs

To make kymographs, we averaged centerline values using a 4-pixel-radius (1.64 *µ*m) circular kernel and plotted them with respect to time. We removed horizontal banding from an image by subtracting a smoothed version of the row means from the image. We obtained lines representing blood packet trajectories from kymograph images by applying a sigma 2 Gaussian blur to the image, using Canny edge detection, and applying the Hough Transform to find lines. We calculated velocities by measuring the average of the slopes of these lines, weighted by the length of the line (Fig. 8 c).

We validated velocities using a custom-made graphical user interface (GUI), which plots the called velocity on top of the kymograph (Fig. 8 d). We would manually determine if the velocity was: Correct (T/F), zero/horizontal (T/F), ‘too slow’, or ‘too fast.; If the velocity call was correct, we retained the called value. If incorrect but the kymograph showed horizontal lines (zero flow), we marked the “Corrected Velocity” as zero. Otherwise, we classified the velocity as “too fast” or “too slow.” We then manually annotated these ‘too fast’ and ‘too slow’ velocities by overlaying the following velocities– 20, 35, 50, 75, 110, 160, 220, 290, 360, 420, 500, 600, 750, 1000, 1500, 2000, 3000, 4000 *µ*m/s–onto the image and choosing the correct slope.

#### 2.5.6 Median Velocities

To normalize for unequal numbers of capillaries in each video when comparing participants, we calculated the median velocity across distinct capillaries for each video. The median of these medians is the median participant velocity. We used medians instead of means because high-velocity values had a larger uncertainty than smaller values.

## 3 Stabilization

To assess stabilization, we analyzed translation values of 1,440,311 frames from 2,253 capillary videos registered using the MOCO plugin which registers each frame in the video to a single reference frame—usually the first—and outputs a log file documenting how much each image was shifted. Aggregating translations from all videos involving 72 participants, we observed that the standard deviation of translation (in pixels) decreased as pressure increased with a Spearman coefficient of -0.231 (Fig. 9 a).

**Fig. 9:**
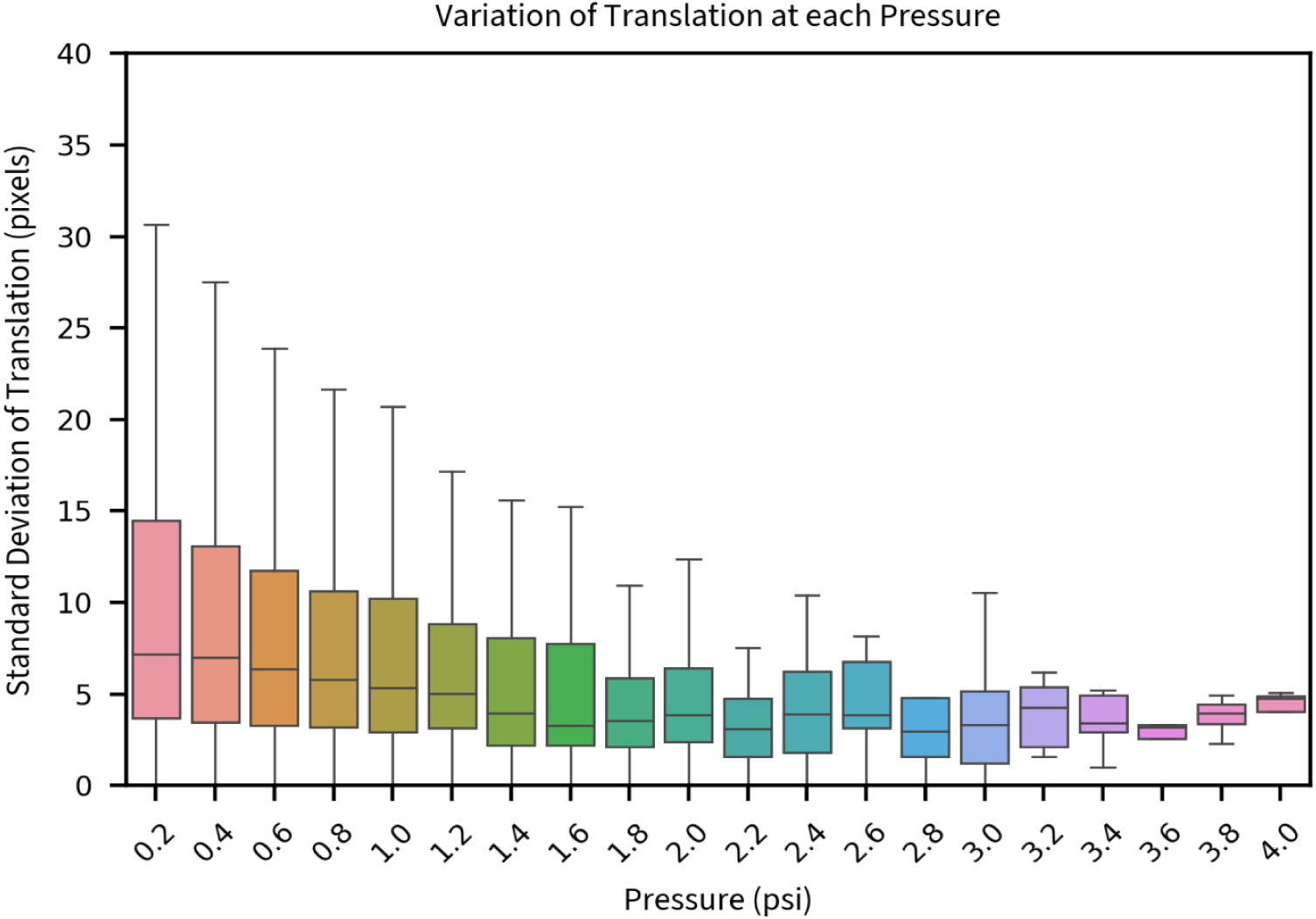
External pressure improves stabilization. Increasing external pressure on the finger decreases the translation necessary to stabilize the video with a Spearman coefficient of -0.231.

## 4 Results

Increasing external pressure on the finger caused a progressive decline in capillary blood cell velocity, ultimately stopping flow at higher pressures (Fig. 10). This shows that studies measuring capillary WBC and RBC velocities must control or calibrate stabilizing pressure to ensure accurate and reproducible results. Using a mixed-effects model to quantify relationships between participants, we saw substantial variation in maximum blood cell velocity across participants (Table 2, ‘Group Variance’) as well as differences in the rate at which velocity declined with increasing pressure (Table 2, ‘Pressure Variance’). Notably, participants with higher baseline blood cell velocities exhibited more gradual reductions in flow under increasing pressure (Table 2, ‘Group × Pressure Covariance’), suggesting that individuals with faster initial flow rates may have better capillary compliance. Future studies should explore whether factors such as age, sex, or cardiovascular health influence these individual differences in capillary reactivity.

**Fig. 10:**
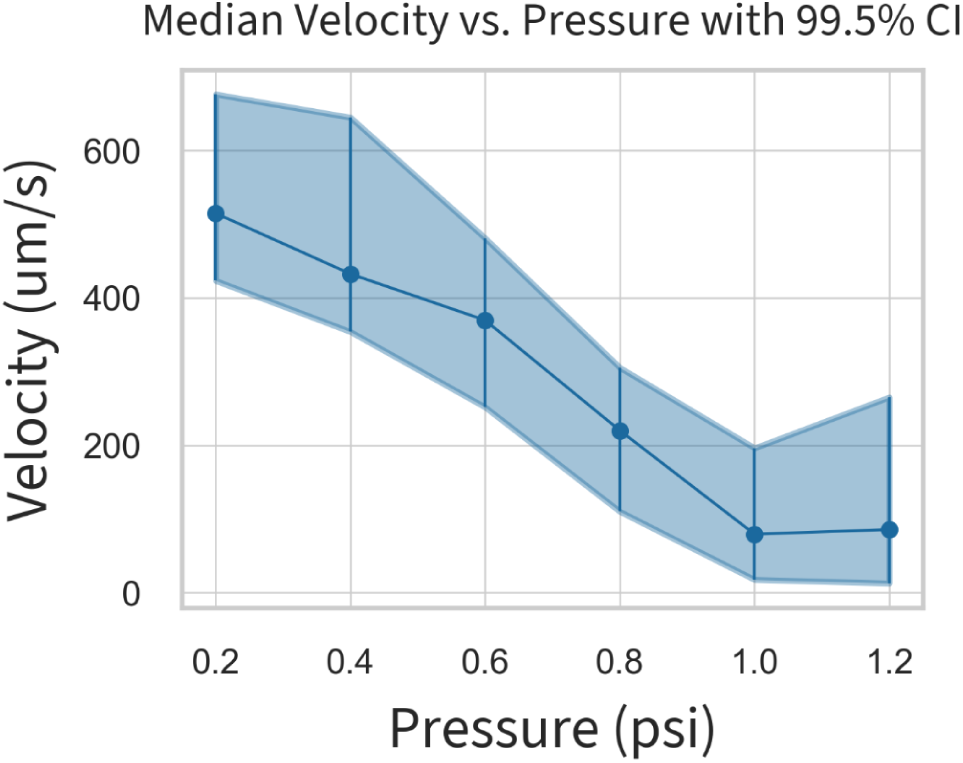
Increasing external pressure decreases blood cell velocity. Median blood cell velocity at each external pressure shows that as external pressure increases, velocity slows and then stops.

**Table 1:** Mixed Linear Model Information.

| Model Characteristic | Value |
| --- | --- |
| Dependent Variable | Log Video Median Velocity |
| Method | REML |
| Number of Observations | 1,826 |
| Number of Groups | 72 |
| Minimum Group Size | 18 |
| Maximum Group Size | 50 |
| Mean Group Size | 25.4 |
| Scale | 3.2294 |
| Log-Likelihood | -3835.6625 |
| Convergence Status | Yes |

**Table 2:**
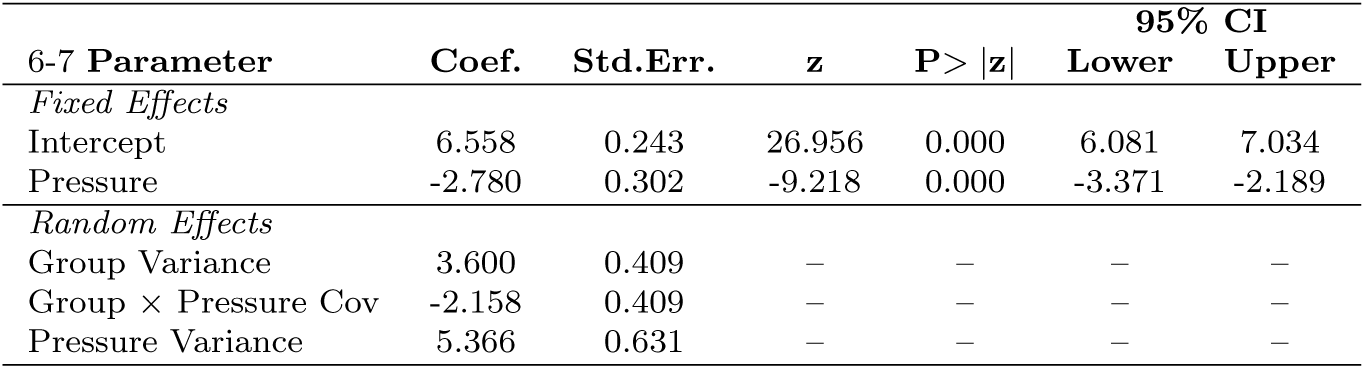
Fixed and Random Effects Estimates.

## 5 Conclusion

We developed an inflatable ‘finger-lock’ that stabilizes nailfold capillaroscopy videos by applying a controlled, constant pressure to the finger, minimizing motion artifacts and enabling precise blood cell velocity measurements. We showed that increasing external pressure on the finger progressively slows RBC and WBC velocities. This highlights the need to standardize or calibrate external pressure for accurate, reproducible capillary flow measurements. Beyond calibration, measuring blood cell velocities enables the *in vivo* assessment of microvascular perfusion and reactivity, which have been shown to precede organ dysfunctions[5–8, 12, 13, 24–27, 53–56], suggesting that skin microvasculature blood flow changes are a reliable method for assessing endothelial function. Our results build on this, with flow responses to pressure varying widely among participants, suggesting that physiological factors such as age, disease status, and cardiovascular health may influence microvascular response to external pressure. Future studies should investigate these relationships to refine capillaroscopy as a new tool for assessing microvascular function. By enabling precise, *in vivo* modulation of microvascular blood flow, the finger-lock provides a new tool for studying capillary reactivity and hemodynamics in real-time, laying the foundation for non-invasive assessment of vascular function, with potential applications in diagnosing and monitoring vascular disorders.

## Data Availability

The data that support the findings of this study are available on request from the corresponding author. The data are not publicly available due to HIPAA restrictions.

## Acknowledgements

We thank G.E. Marti for optics and statistics discussions. We thank J. Lee, T. Chen and S. Cofer for helpful discussions. We thank Hongquan Li for optics and software discussions. We thank K. Medill for electrical circuit input, feedback, and helpful discussions. We thank N. Kozak and A. Marsden for fluid-flow discussions. Funding: This work is supported by the Chan Zuckerberg Biohub. M.L.F. is supported by a National Science Foundation Graduate Research Fellowship, and the Knight-Hennessy Scholars Program.

## Funding

- Knight-Hennessy Scholars Program
- NSF GRFP Scholarship
- CZI Biohub grant: 1197775-1GWMUM
- Stanford Diabetes Research Center (SDRC) grant number: P30DK116074

### Ethical approval

1. Approval: All experimental methods were approved by Stanford institutional review board (IRB) 6 in protocol number 64537.
2. Accordance: All methods were carried out as written in the IRB protocol 64537.
3. Informed consent: Informed consent was obtained from all participants.

### Code availability

All code is available at: https://github.com/gt8mar/capillary-flow

### Data availability

The data that support the findings of this study are available from the corresponding author upon reasonable request and HIPAA approval.

### Author contribution

M.L.F and S.R.Q. conceptualized the study. M.L.F. and S.R.Q. designed the study. M.L.F., S.R.Q., and D.N.C. wrote the IRB protocol. G.R. wrote the capillary-naming pipeline. M.L.F. wrote all other parts of the data processing and analysis pipeline. M.L.F, G.R, J.H.L, S.R.Q., and D.R.C recruited all participants. M.L.F., G.R., and J.H.L. collected, processed, and performed quality control on all data. M.L.F. performed all analyses. M.L.F. and S.R.Q. wrote the manuscript. All authors revised the manuscript and approved it for publication.

